# Age-dependent and Independent Symptoms and Comorbidities Predictive of COVID-19 Hospitalization

**DOI:** 10.1101/2020.08.14.20170365

**Authors:** Yingxiang Huang, Dina Radenkovic, Kevin Perez, Kari Nadeau, Eric Verdin, David Furman

**Affiliations:** Buck Institute for Research on Aging, Novato, CA 94945, USA; Guy’s & St Thomas’ NHS Foundation Trust and King’s College London, Westminster Bridge Road, London SE1 7EH, UK; Stanford 1000 Immunomes Project, Stanford University School of Medicine, Stanford, California, 94305, USA; Sean N. Parker Center at Stanford University, Division of Pulmonary, Allergy, and Critical Care, Department of Medicine, Stanford, California, 94305, USA; Austral Institute for Applied Artificial Intelligence, Institute for Research in Translational Medicine (IIMT), Universidad Austral, CONICET, Pilar, Buenos Aires, B1630FHB, Argentina

## Abstract

The coronavirus disease 2019 (COVID-19) pandemic, caused by Severe Acute Respiratory Syndrome (SARS)-CoV-2, continues to burden medical institutions around the world by increasing total hospitalization and Intensive Care Unit (ICU) admissions^1–9^ A better understanding of symptoms, comorbidities and medication used for preexisting conditions in patients with COVID-19 could help healthcare workers identify patients at increased risk of developing more severe disease^10,11^. Here, we have used self-reported data (symptoms, medications and comorbidities) from more than 3 million users from the *COVID-19 Symptom Tracker* app^12^ to identify previously reported and novel features predictive of patients being admitted in a hospital setting. Despite previously reported association between age and more severe disease phenotypes^13–18^, we found that patient’s age, sex and ethnic group were minimally predictive when compared to patient’s symptoms and comorbidities. The most important variables selected by our predictive algorithm were fever, the use of immunosuppressant medication, mobility aid, shortness of breath and fatigue. It is anticipated that early administration of preventative measures in COVID-19 positive patients (COVID+) who exhibit a high risk of hospitalization signature may prevent severe disease progression.

## Main

The *COVID-19 Symptom Tracker* is a smartphone app where individuals from the (United Kingdom) UK and (United States) US can submit their symptoms daily^19–22^ A total of 3,485,804 users have signed up for the app as of July 1st, 2020^12^. A user can have multiple entries spanning multiple days recording features such as symptoms, comorbidities, medication for pre-existing conditions, and demographics. The features we used in all subsequent models are listed in Table 1. All features were binary except for age and BMI, which were continuous; and shortness of breath (SOB), fatigue, race, and gender, which were categorical. For the study cohort, we extracted all users who tested positive for COVID-19 (n = 10,948). Of those COVID+ users, some cases were severe enough to require them to visit the hospital while others managed their disease at home (Fig. S1). We used comorbidities, demographics, and symptoms to predict patients’ admission to a hospital setting. To do so, we first divided the COVID+ patients into two groups: (A) negative for hospitalization, including COVID+ patients who were strictly at home without ever having to be admitted to a hospital setting (n = 10,413) and (B) positive for hospitalization, including COVID+ users who reported being admitted to the hospital (n = 535). The average age of group A was 40.2 (Standard Deviation: 13.6) compared to 47.8 (Standard Deviation: 18.8) for group B. For group A, we used comorbidities, demographics, and symptoms recorded in the patient’s last entry, and for group B, we used features recorded one entry prior to the entry where the patient indicates admission to a hospital setting (scenario 1) (see Methods). We also analyzed the data considering whether a patient ever reported a given symptom along with comorbidities, demographics, and pre-existing medications (scenario 2) with similar results to those of scenario 1.

**Table 1.**
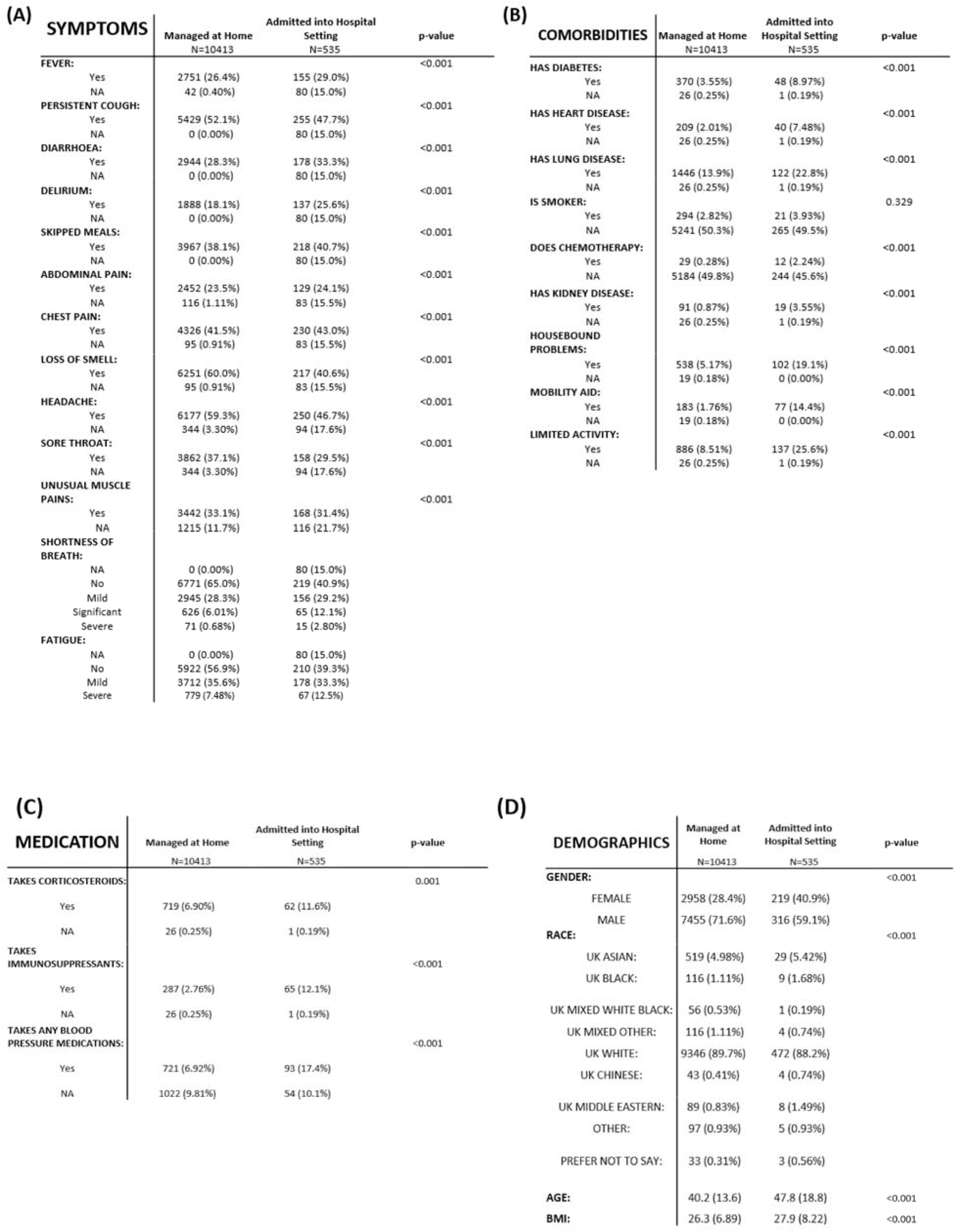
Features used in Elastic Net Model. Features of symptoms, medication history, comorbidities, and demographics investigated in relations to whether a user was admitted to a hospital setting. All features were binary except for age and BMI, which were continuous, and shortness of breath, fatigue, race, and gender, which were categorical. For each feature, NA indicates not available/missing data.

We performed an Elastic Net regularized regression to analyze the predictive performance of the features and used LASSO regularization to select for the most important features for the prediction of patient’s admission to a hospital setting. The dataset was divided into training and test sets (ratio: 70:30). Since patients often neglect to report all available fields, we used the multiple imputations method to account for missing values, a standard procedure to predict missing data using all other features (besides the outcome) that are not missing^23–25^. Since the number of patients in group A was considerably larger than in group B (class imbalance) both undersampling of the majority cases and oversampling of minority cases was utilized to achieve a balanced training set (see Methods). Using cross-validation on the training set, parameters are tuned for the Elastic Net Regression, producing the best predictive performance and the most parsimonious number of features. We were able to predict patient hospitalization with relatively good accuracy (cross-validated area under the receiver operating curve (cvAUC) for the training set at the optimal parameters was 0.77) (Fig. 1A). Using the features selected by this analysis (Fig. 1B) for the prediction of hospitalization on the test set, a similar accuracy was obtained (cvAUC = 78%) (Fig. 1C). The most important variables of this signature selected by our predictive algorithm were fever, the use of immunosuppressant medication, mobility aid, shortness of breath and fatigue. Age had a relatively small regression coefficient indicating that pre-existing clinical conditions and symptom presentation are much stronger predictors of hospitalization. Unexpectedly, the body mass index (BMI) was not selected as a significant predictor. Finally, the female gender was negatively associated with hospitalization.

**Figure 1.**
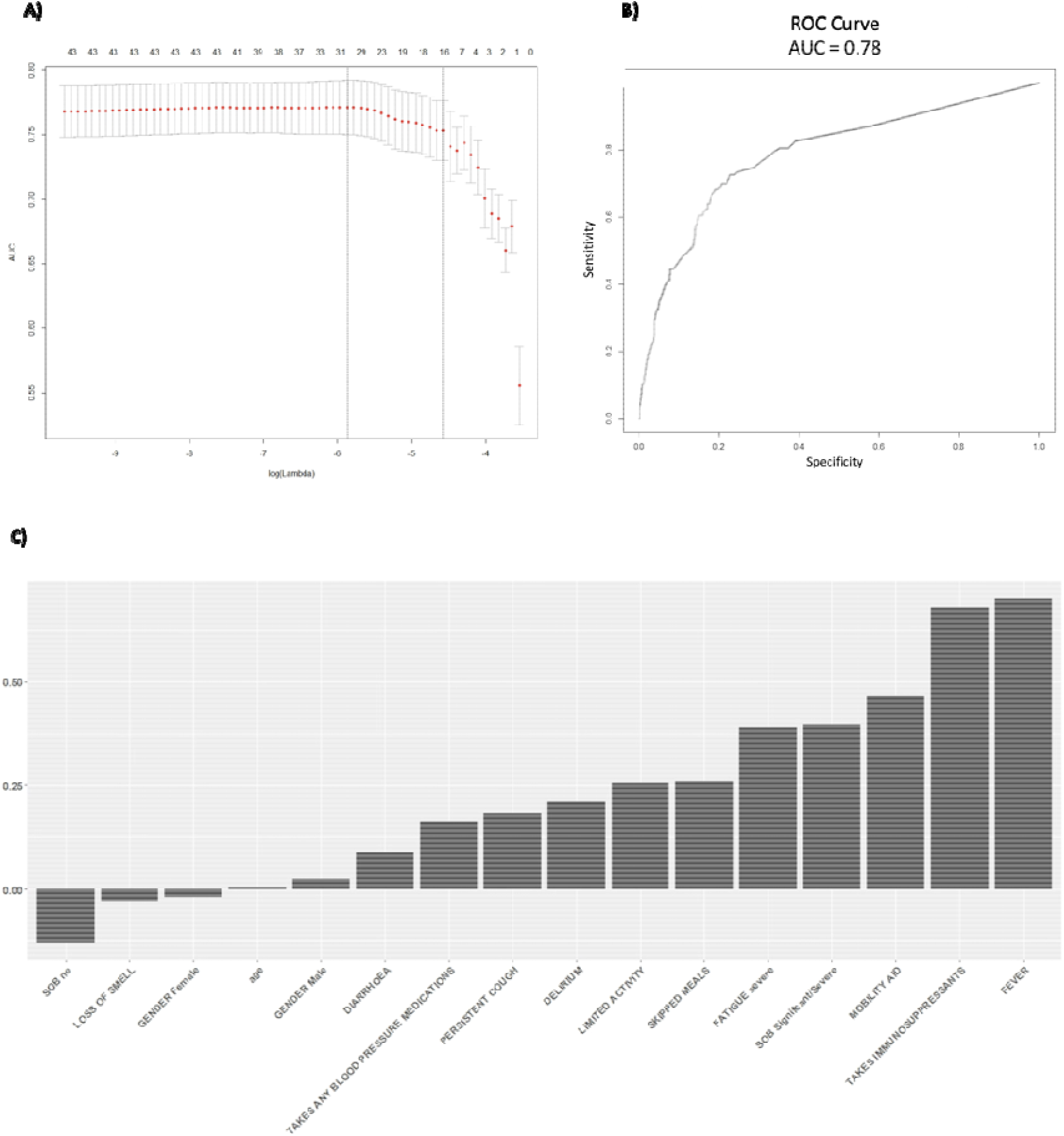
Elastic Net Regression predictive performance and selected variables. We used Elastic Net Regression where outcome of being admitted in a hospital setting or not was regressed on features in **Table 1**. **(A)** The performance in terms of crossvalidation area under the Reciever Operating Curve (AUC) for validated Elastic Net Regression on the training set across different values of lambda. **(B)** The AUC of the trained Elastic Net model applied on a holdout test dataset. **(C)** The most important features selected by the Elastic Net model. Negative coefficients indicate a negative association with outcome and vice versa.

We next estimated the odds ratio from logistic regression for each feature where the outcome (being admitted in a hospital setting) was regressed onto all features (Fig. S4). The most important features are consistent with the Elastic Net results. Elastic Net Regression was also applied to scenario 2. The prediction performance is comparable to scenario 1, and the selected features were also very similar (Fig. S3). The modeling from logistic regression and Elastic Net regression using scenario 1 and 2 all selected similar features that are predictive of the outcome, lending robustness to the results.

To understand the age effects better given that it has small significance in predicting the outcome, we analyzed the association between age and the other features selected. We conducted an experiment where we divided all the COVID+ users into three age groups, young, middle age, and old. Running univariate logistic regression where the outcome of being admitted to a hospital setting is regressed onto each feature selected by the Elastic Net model shows that the coefficients of the features do not vary substantially between age groups (Fig. S7). Such results suggest that the features’ association to the outcome is not dependent on age.

To better understand the fluctuations in the symptoms selected by the Elastic Net model, we then analyzed the eight symptoms in a longitudinal manner. We examined a window of 20 days before the patient goes to the hospital (for positive cases), and 20 days before the last entry (for negative cases) (Fig. 2). For each day, we estimated the frequency of each symptom for the positive and negative groups. Day 0 for the positive group corresponds to the day when the patient was admitted to a hospital setting, and day 0 for the negative group corresponds to the last patient’s entry. Fig. 2A shows positive and negative groups of binary variables. Fig. 2B shows categorical variables of fatigue and SOB for the positive group, and Fig. 2C shows fatigue and SOB for the negative group. A linear regression line is superimposed for each group where the frequency is regressed on the days. Slope and intercepts are shown for comparison and their significance is evaluated using the likelihood ratio test (Fig. S7). All differences between the two groups were significant except for mild fatigue. The slopes of the positive group were steeper than in the negative group in all the symptoms except for diarrhea, which indicates that the positive group increased in frequency of symptoms that are indicative of severe COVID-19 cases as the disease progressed while the frequency of the symptoms for the negative group stayed relatively stable. Not surprisingly, all the intercepts for the positive group are higher than the negative group except for mild fatigue, further indicating that there are higher frequencies of COVID-19 related symptoms in users who were admitted to a hospital setting.

**Figure 2.**
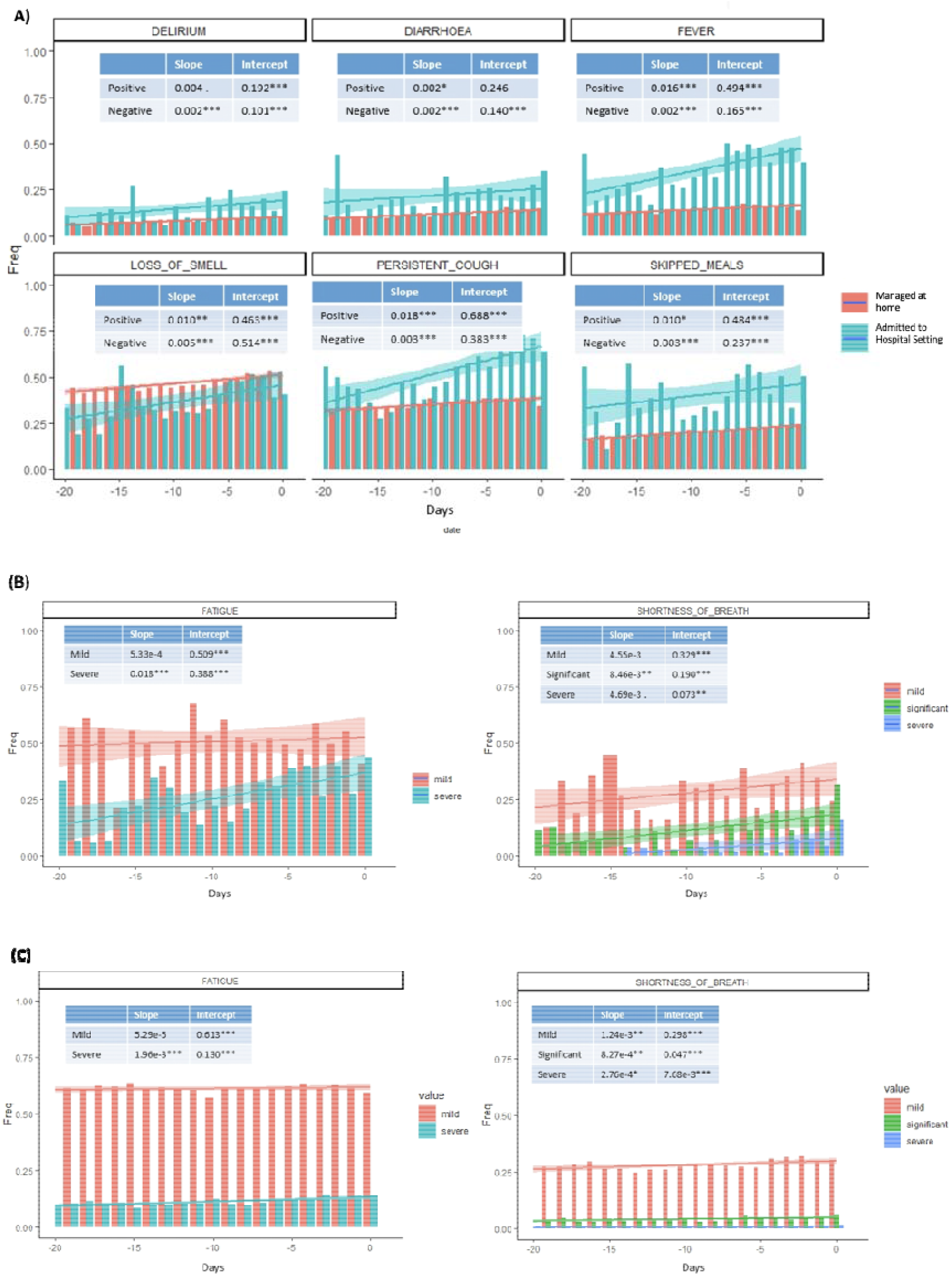
Trajectory analysis of features selected from Elastic Net. We analyzed all Elastic Net Regression selected symptoms for a 20 days window where day 0 for the positive group is the 20 days before the user goes to the hospital and day 0 for the negative group is the 20 days before the user’s last entry. Frequency of users having each feature for each day were plotted. A linear regression line where the frequency is regressed onto the days is plotted. The slopes and intercepts are labeled. **(A)** Binary features of both positive and negative groups are plotted. **(B)** Categorical features of fatigue and shortness of breath are plotted for the positive group. **(C)** Categorical features of fatigue and shortness of breath are plotted for the negative group.

SARS-CoV-2 has been shown to cause more severe diseases in older adults ^26^ Even though age was not a major contributor to the prediction of COVID-19 related hospitalization, we explored whether age was associated with other features selected by the model. In conjunction, we also examined other demographic variables, such as race, BMI and gender. We conducted multivariate logistic regression models where each of the features selected by the Elastic Net model was regressed on the demographic variables analyzed (Fig. 3). Age was associated with 10/13 of the predictive features (*P* < 0.01). The most age-correlated features were mobility aid, limited activity, blood pressure medication and immunosuppressant medication use. This indicates that age-related phenotypes in this cohort are associated with hospitalization due to COVID-19. This emphasizes the fact despite age, any population that expresses the features selected from our model could be susceptible to a more severe form of COVID-19. Understanding vulnerable young populations that make them biologically older than their chronological age and exhibit features that are generally associated with the older population could help identify susceptible young populations.

**Figure 3.**
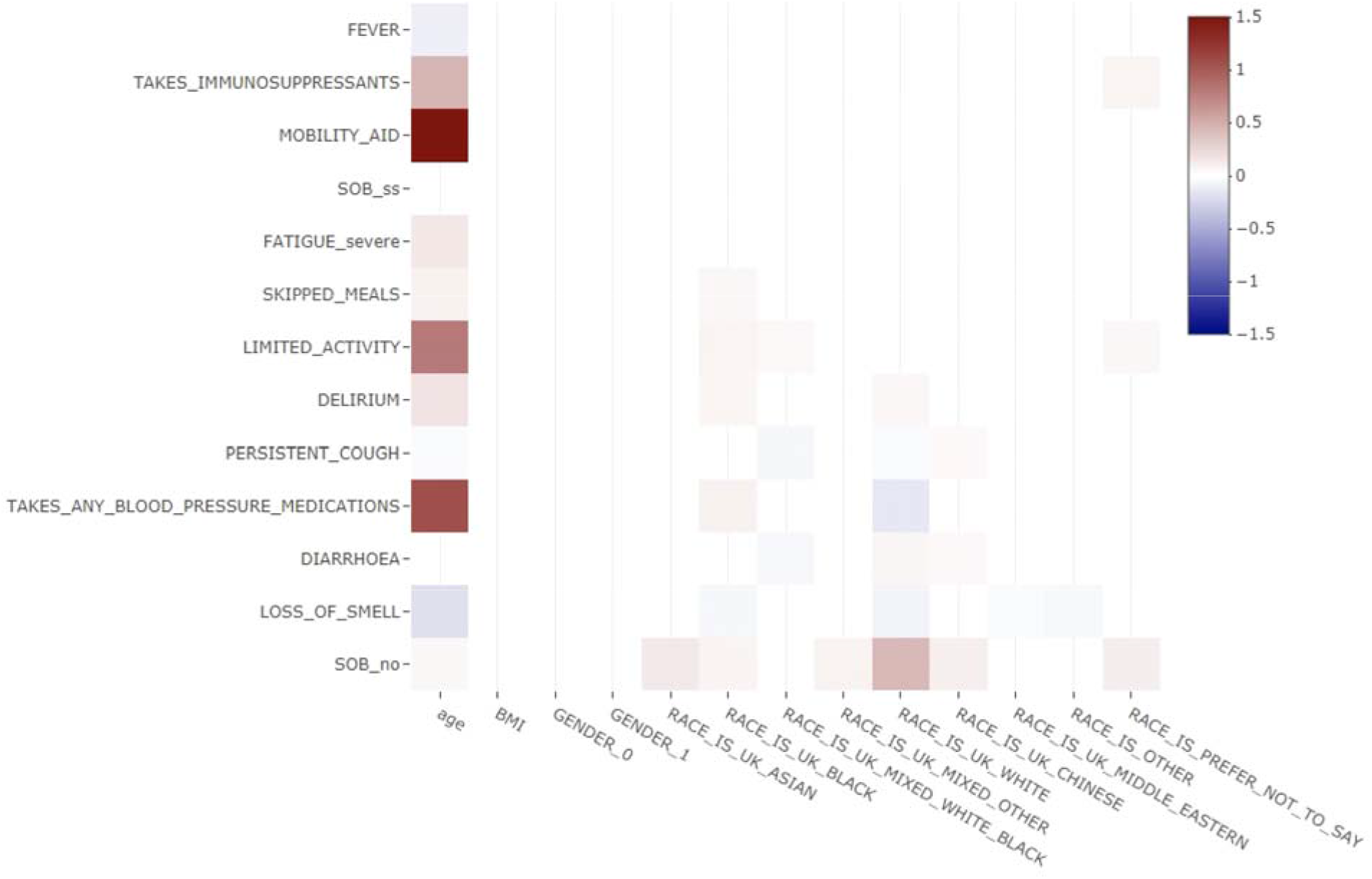
Demographics correlation to Elastic Net Regression selected features. Multivariate logistic regression where each Elastic Net Regression selected feature is regressed onto demographic information such as age, BMI, gender, and race. Coefficients are plotted in a heatmap. Only statistically significant associations are plotted. Age has significant but weak association with many selected features. Users identified as black ethnicity in the UK have many positive associations with high coefficients.

In addition to age, being of black ethnicity was associated with a number of features selected by the Elastic Net such as a high frequency of delirium, limited activity, and blood pressure medications usage. However, whether this is associated with social-economical status or an innate biological difference in people of African descent need further investigation. The gender feature was a predictor of hospitalization (Fig. 1C) but was not significantly correlated with any of the predictive features suggesting that the sex of an individual affects other aspects of disease severity not evaluated in this study.

A relatively small effect of the loss of smell feature associated with mild disease outcomes (Fig. 1C) have also been reported in recent studies ^27,28^ However, we also show that this feature is age and race associated. The female gender also had a negative correlation with hospitalization consistent with recent findings in large populations ^29,30^ In our data, gender did not correlate strongly with any features, indicating that there may be factors other than comorbidities or symptoms that make females have a better prognosis. Underlying immunological differences in females ^31–34^ could lead to the mounting of a better immune response that could neutralize the virus more efficiently than in men.

From our analyses, we have found features that are predictive of people having severe enough COVID-19 disease to be admitted into a hospital setting. However, there are some features where additional research would help elucidate the mechanism behind their correlation. For example, immunosuppressant use was a major predictor of patient hospitalization and from the data we cannot investigate whether patients taking these drugs are more prone to severe COVID-19 because underlying autoimmune/auto-inflammatory disease or because of the direct effect of the drug on the suppression of the inflammatory response. If the latter was true immunosuppressant use should ameliorate severity since severe disease phenotypes are initiated by a cytokine storm^35^ which could be attenuated by the use of immunosuppressant medication.

Besides the need for additional research into the mechanism behind some of the features associated with more severe disease state, time is an important variable that is not explored in depth in this paper. A Cox survival analysis would be informative, however, the start time of each user is inconsistent and thus, the application of Cox survival is inappropriate. Some users’ first entries already indicate testing positive for COVID-19 with symptoms suggesting that they are already in the midst of the disease course, while others slowly develop symptoms and test positive for COVID-19 later in time.

Age has been shown to be important in the severity of COVID-19^13^. In our results, age shows a slight positive correlation with being admitted in a hospital setting. The difference between the average age of those who were admitted to a hospital and those who did not was relatively small, consistent with age not being a strong predictor. It is possible that the older population was less likely to use a smartphone app, leading to under representations of the sick older population. The fact that age-associated variables outperform age in the prediction of patient hospitalization indicates that biological age or immunological age^36,37^ could be appropriate measures in assessing an individual’s prognosis.

In conclusion, we identify age-dependent and independent sets of symptoms and comorbidities predictive of COVID-19 patient hospitalization. Our analyses show features that predict disease severity in advance and this can be utilized to inform severe cases of COVID-19 even in younger individuals who may not be labeled as high risk. Continued rise in the number of cases, as societies struggle to balance reopening the economy and ‘flattening the curve’, places an enormous burden on healthcare systems around the world. Knowing the signs of possible severe cases like the ones derived in this study could help healthcare systems devote resources to intervening in potentially severe cases before they become costly to manage.

## Methods

### Study Cohort

Of all the users who signed up for the *Tracker* app, we extracted all users who have indicated testing positive for COVID-19 from March 24, 2020 to June 23, 2020 (Fig. S1). United States users were excluded from the study to maintain homogeneity of the study cohort, reducing potential noise. Users who did not enter values for more than 90% of variables were excluded. It is extremely difficult to impute the missing values and derive any meaningful analysis from such users.

### Outcomes and features of Scenario 1 and 2

From the study cohort, the outcomes or dependent variable that we are interested in is whether a user from the *Tracker* app is admitted to a hospital setting in any capacity or not. Since the users can enter their symptoms everyday, there are many time points we can use as features. For what we call scenario 1, for users who were admitted to a hospital setting, we used the time point right before a user indicated he/she is in the hospital and the features at that time point for analysis. For users who were always at home, we used the last time point and the features at that time point for analysis (Fig. S2A). In what we call scenario 2, for users who were admitted to a hospital setting, if a user indicated that he/she had a feature in any of his/her entire entries before the day of being admitted in a hospital setting, we labeled that feature as positive for that user. For users who were always at home, if he/she had a feature for his/her entire entry log, we labeled that feature as positive for that user (Fig. S2B). Using such methods only apply to symptoms since they can change everyday and not to comorbidities, pre-existing medication use, or demographics.

### Imputation

Multiple imputations were used to impute missing values. Instead of imputing the missing value with a single value, multiple imputations repeatedly samples the data *n* times and impute the missing values *n* times using different methods for different data type. We used predictive mean matching for numerical data (age, BMI), polytomous regression for unordered categorical data (gender, race), proportional odds model for ordered categorical data (fatigue, SOB), and logistic regression for binary data (all other features). The variables for the logistic regression would be all other independent variables while the outcome would be the missing variable. The most stringent process would only impute the training set, but there are not enough complete instances to have both positive and negative cases, therefore, we imputed training and testing together. To account for bias, when creating the test set we assessed the pattern of missingness and sampled each pattern so that the test set is representative of all missingness patterns. Multiple imputations produce *n* imputations, and we pool *n* imputed matrices together to form a larger training set.

Some variables had a large percentage of missing values as seen in Fig. S3B. A comparison of imputed distribution to the original distribution indicates that some variables would produce a wide range of distribution from one imputation to another that is too different from the original distribution (Fig. S3). Therefore, those variables are removed from the datasets.

### Data Balanceness

Class imbalance is an issue given that users who specified they are in the hospital is 1.5% of the total entries. To balance out the training set so that Elastic Net regularization does not bias toward negative cases, we oversampled the positive cases and undersampled the negative cases until the number of positive and negative cases are equal.

### Elastic Net Regularization

Two parameters can be tuned in Elastic Net, alpha and lambda. Alpha is the mixing parameter indicating how much lasso regularization and ridge regularization should contribute to the model. Lambda is the amount of shrinkage or regularization the model should apply as a whole. A series of alpha is used in each cross-validation of lambda. The alpha that produces the highest AUROC at the minimum lambda is chosen. For scenario 1, the alpha is 0.1. Two common lambdas are generally used, the lambda that gives the best performance (lambda.min) or the lambda with the fewest features selected and is within one standard error of the best performing lambda (lambda. 1se). We used lambda. 1se because it is the most generalizable model, avoiding overfitting and selecting the most salient variables.

### Likelihood Ratio Test

The likelihood ratio test was used to compare whether there are statistically significant differences between the slopes of positive and negative cases in the trajectory analysis. Linear regression was used to quantify the association between days and frequency of each selected symptom in positive and negative cases. The likelihood ratio test was used to compare the linear regression model where the frequency of the feature was the independent variable and the linear regression model where the frequency of the feature and whether cases are positive or negative were the independent variables. The null hypothesis is that a linear model with only frequency of the feature as the independent variable is the superior model, the alternative hypothesis is that the superior model is the model with frequency of features and whether cases are positive or negative are independent variables. Rejection of the null hypothesis suggests that knowing positive or negative cases predicts better frequency, therefore the positive and negative cases are statistically different.

## Data Availability

Data can be requested for access through The Health Data Research Hub for Respiratory Health, in partnership with SAIL Databank.

## Supplementary

**Supplementary Figure 1.**
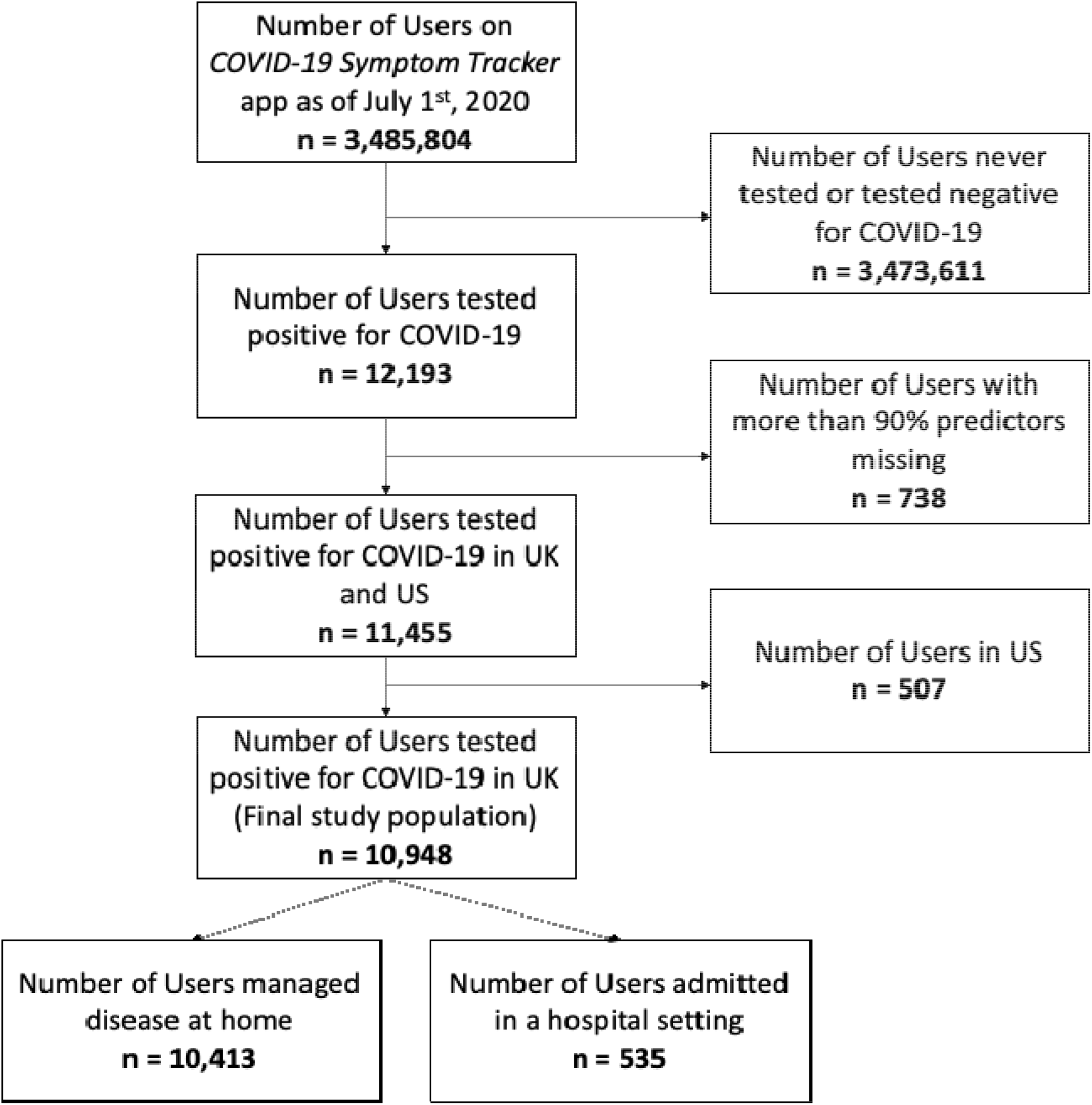
Diagram of cohort with inclusion and exclusion critiera. Only Users tested positive for COVID-19 were included. Users with too many predictors missing were excluded.

**Supplementary Figure 2.**
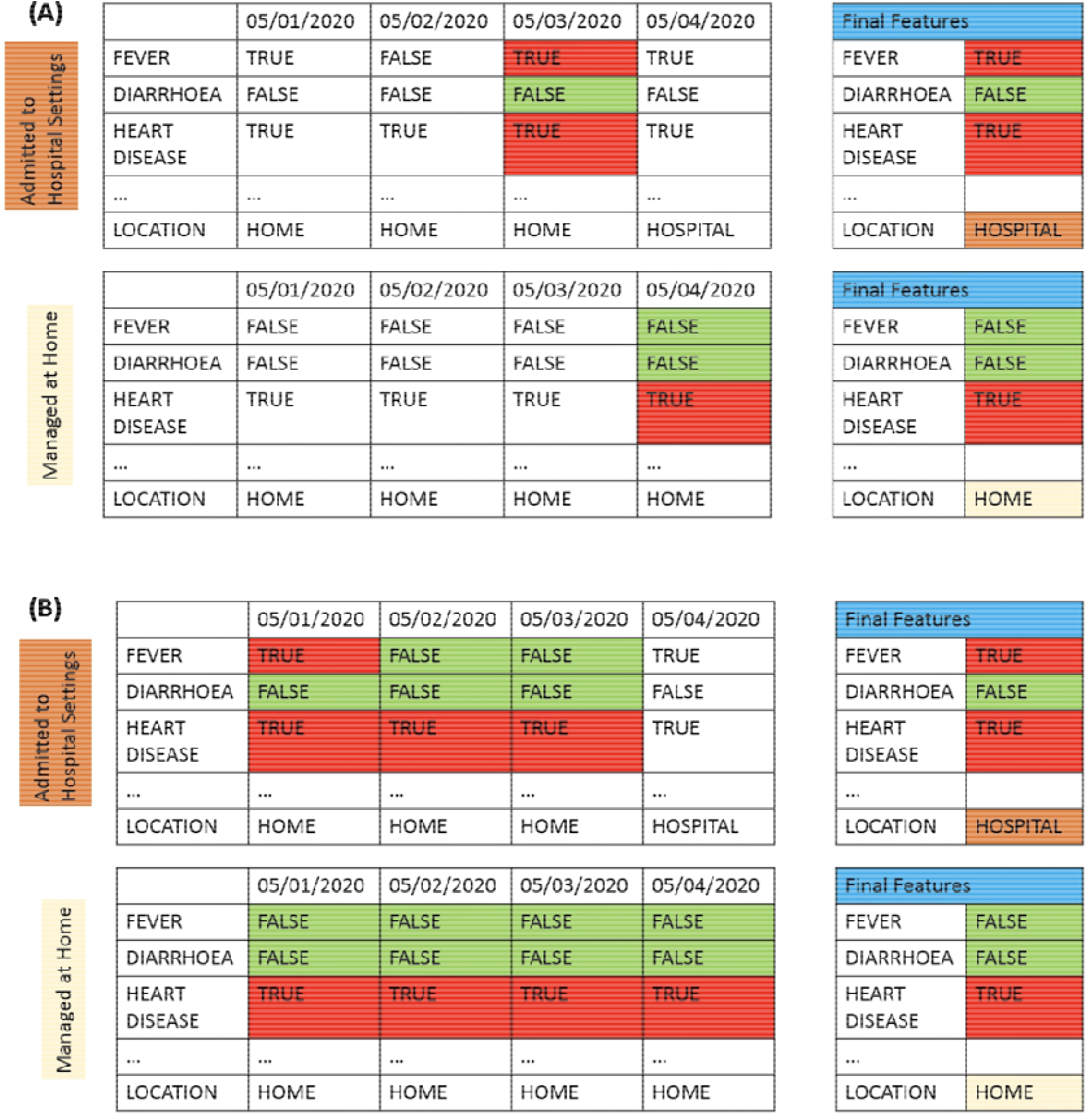
Usage of the features. (**A**) For users who were admitted to a hospital setting, we used the time point right before a user indicated he/she is in the hospital and the features at that time point for analysis. For users who were always at home, we used the last time point and the features at that time point for analysis (**B**) For users who were admitted to a hospital setting, if a user indicated that he/she had a feature in any of his/her entire entries before the day of being admitted in a hospital setting, we labeled that feature as positive for that user. For users who were always at home, if he/she had a feature for his/her entire entry log, we labeled that feature as positive for that user. Such methods only apply to symptoms since they can change everyday and not to comorbidities, pre-existing medication use, or demographics.

**Supplementary Figure 3:**
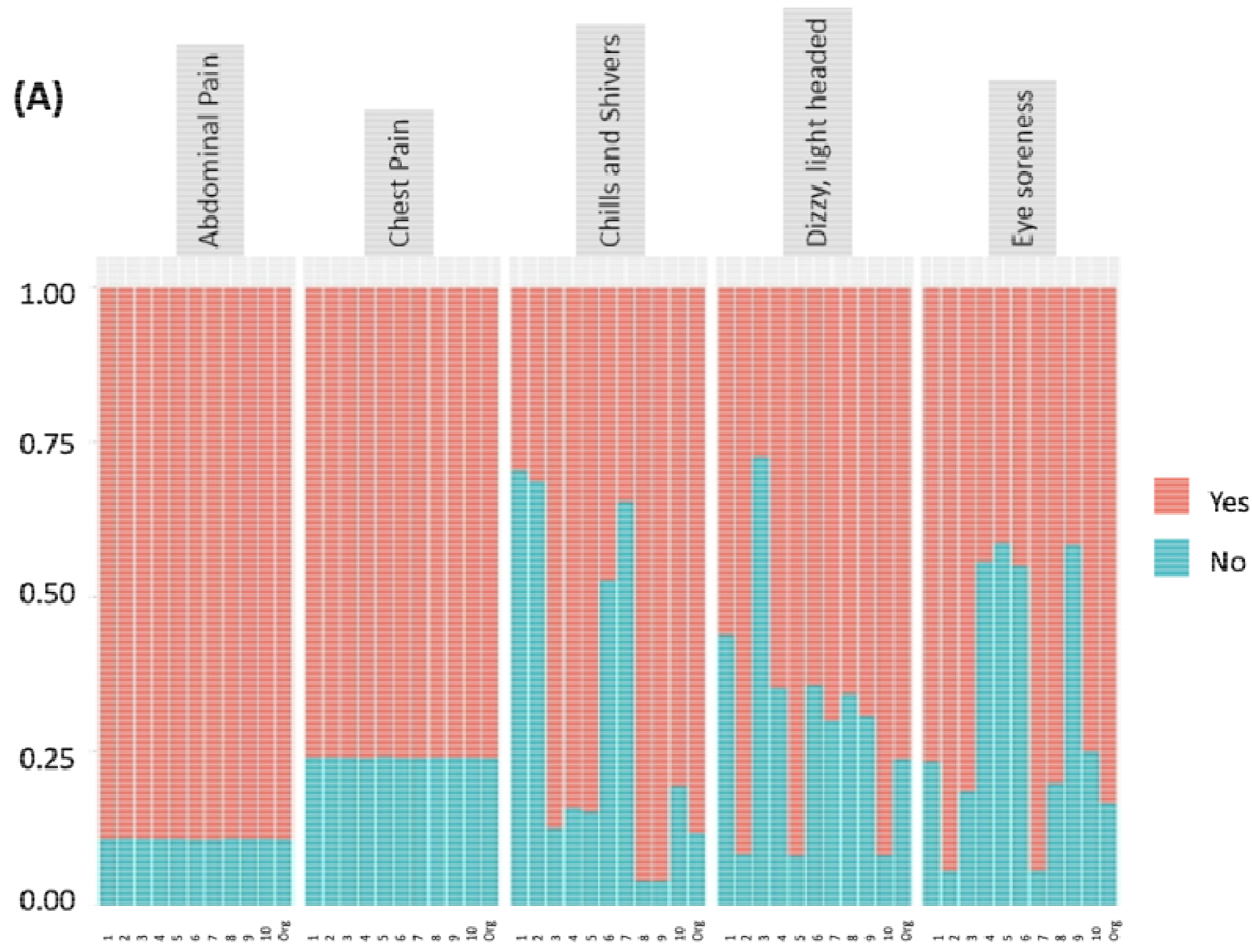
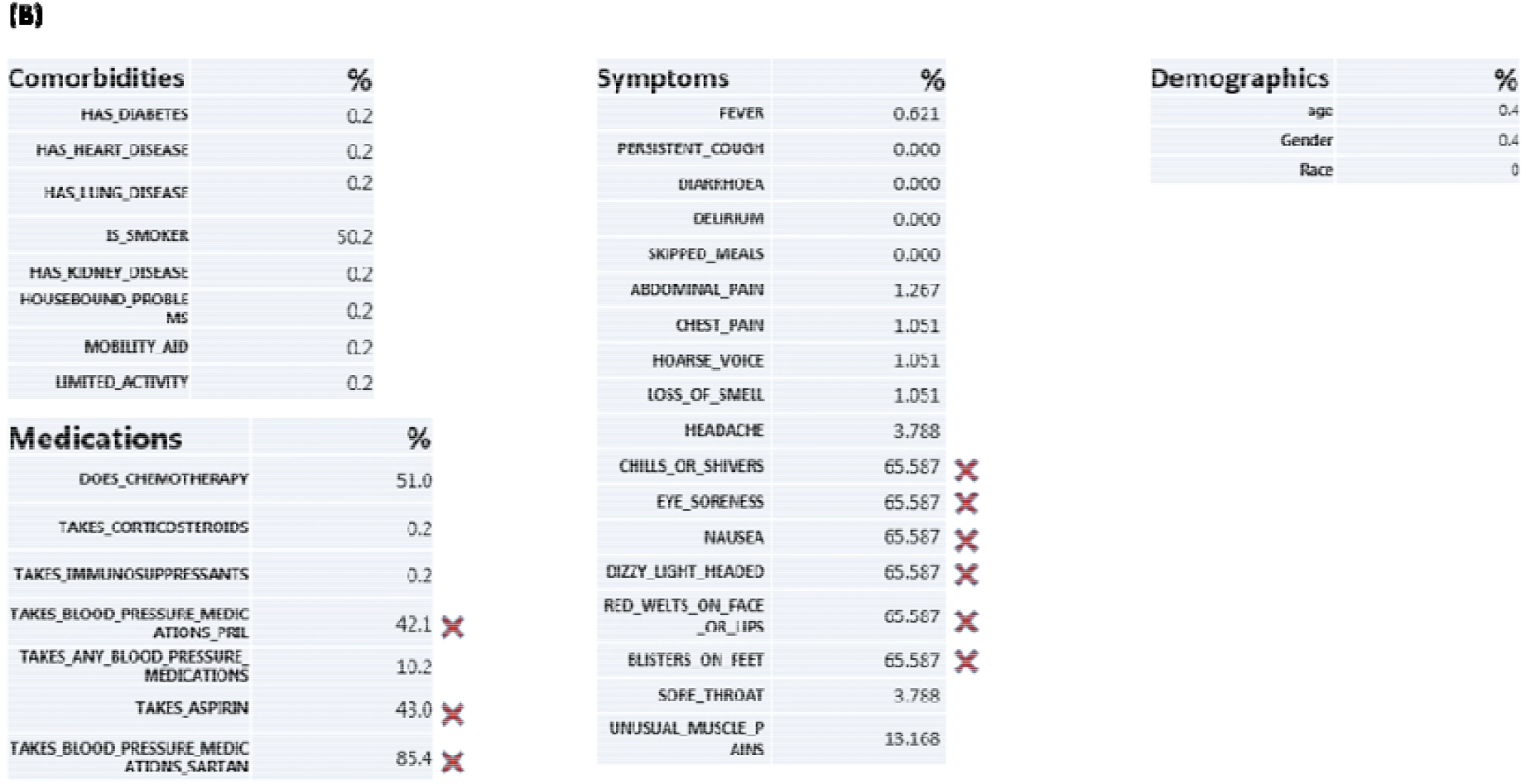
Multiple Imputation of missing values. An example of the original distribution of features and imputed distribution of those features is shown in (**A**). The last column of each feature group labeled ‘org’ is the original distribution without imputation. Labels 1-10 are the ten different distributions after multiple imputations of the missing values. Some features, ‘Abdominal pain’ and ‘Chest pain’ in this example are able to retain the original distribution after multiple imputations. Other features had a wide range of distributions that were wildly different from the original, indicating the multiple imputations for these features were not suitable. Those features were removed from the original dataset. The features removed are labeled with a red cross in (**B**). The percentage missing is shown for each feature.

**Supplementary Figure 4:**
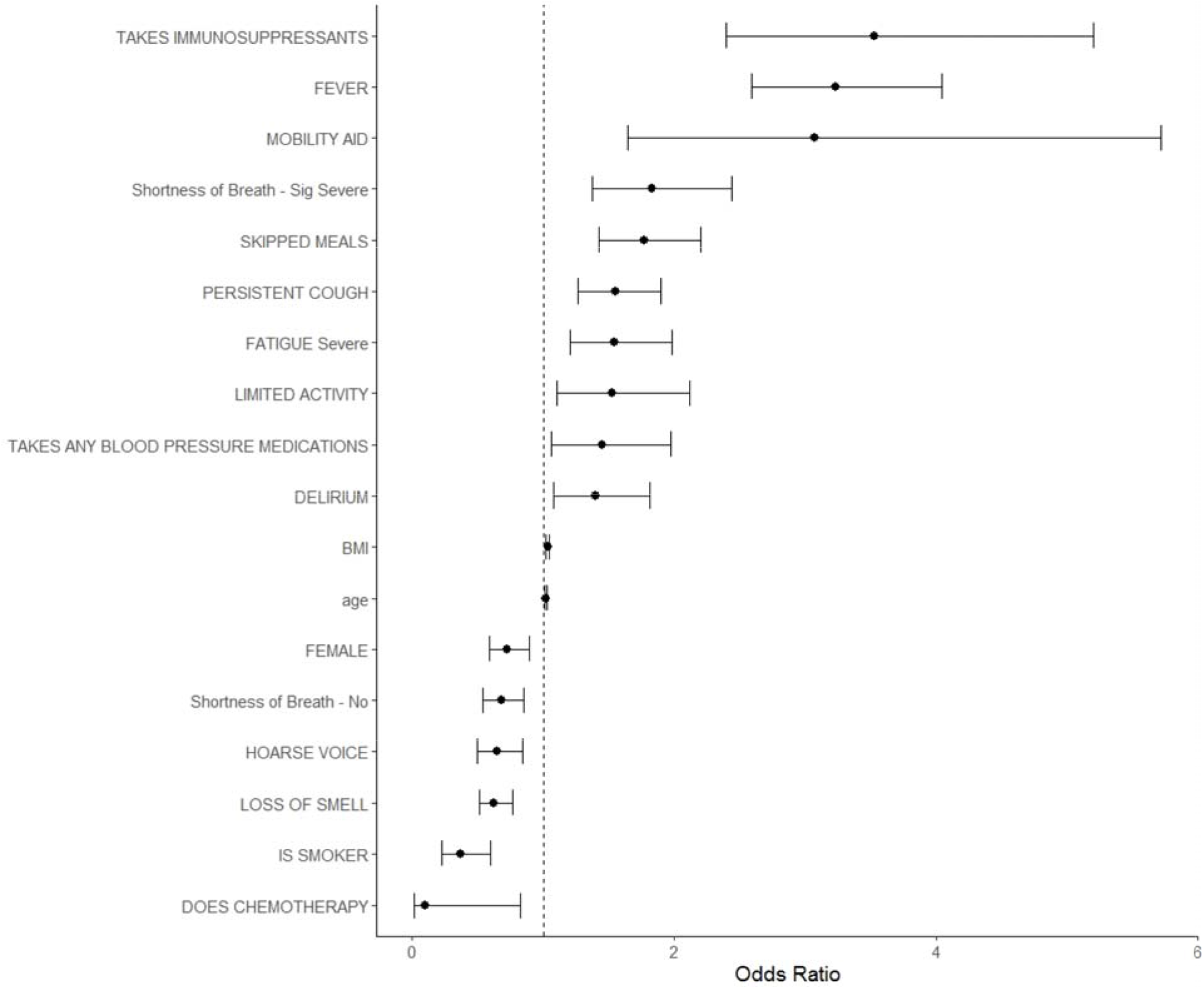
Estimated Odds Ratios for each potential risk factor from a logistic regression model. Error bars represent 95% confidence interval for the odds ratio. All odds ratios are adjusted for all other factors listed. Only Significant features are shown.

**Supplementary Figure 5.**
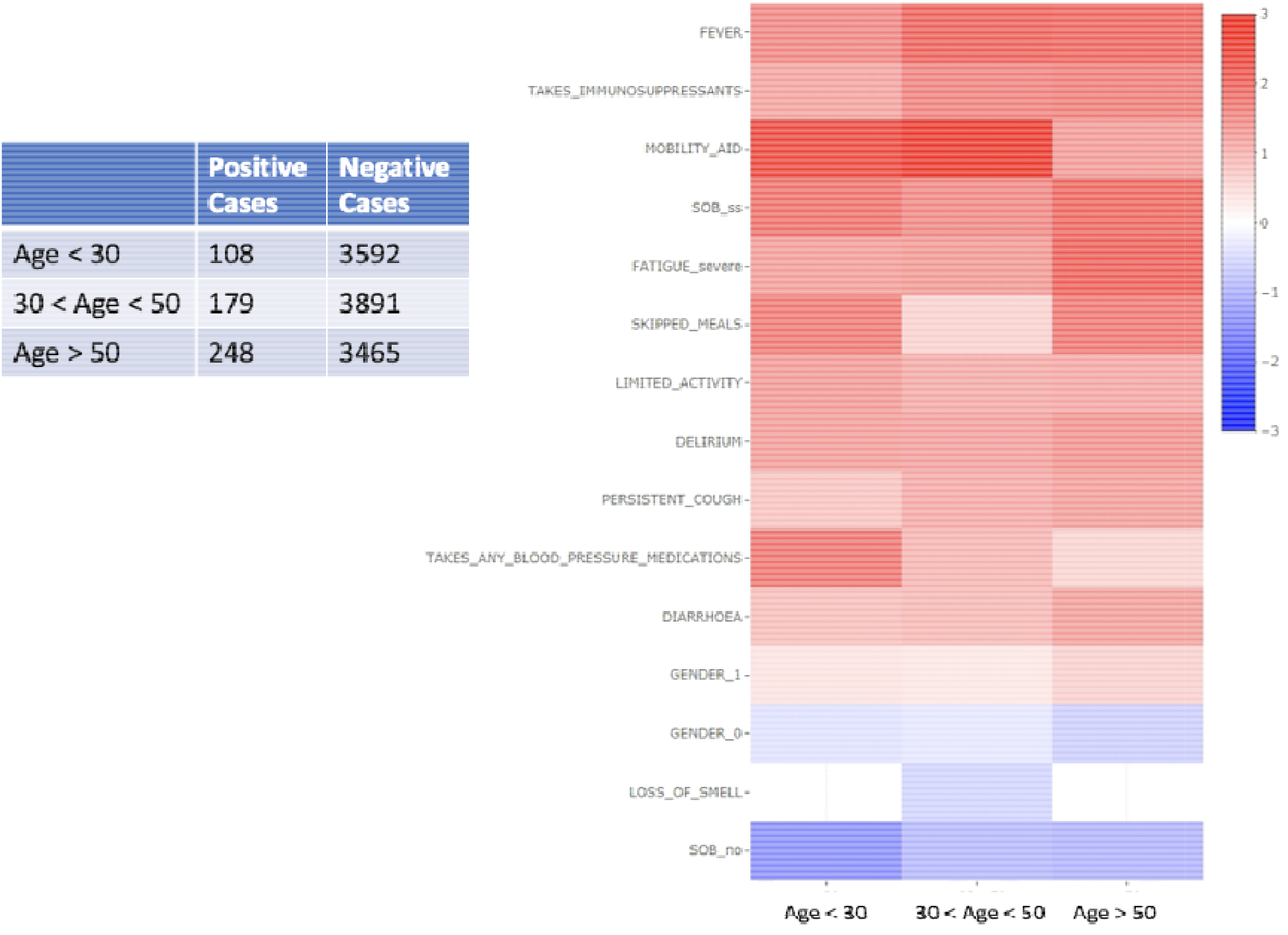
Univariate Logistic Regression of young, middle age, and old age groups. All the COVID+ users were divided into three groups of young, middle age, and old age groups. The number of positive cases (admitted in a hospital setting) and the number of negative cases (stayed home) are shown. The outcome of whether a user was admitted to the hospital was regressed onto each of the features selected by the Elastic Net Regression. The coefficients for each feature for each age group is plotted. Only significant ones are colored. The three groups have similar patterns of expression in the features selected.

**Supplementary Figure 6:**
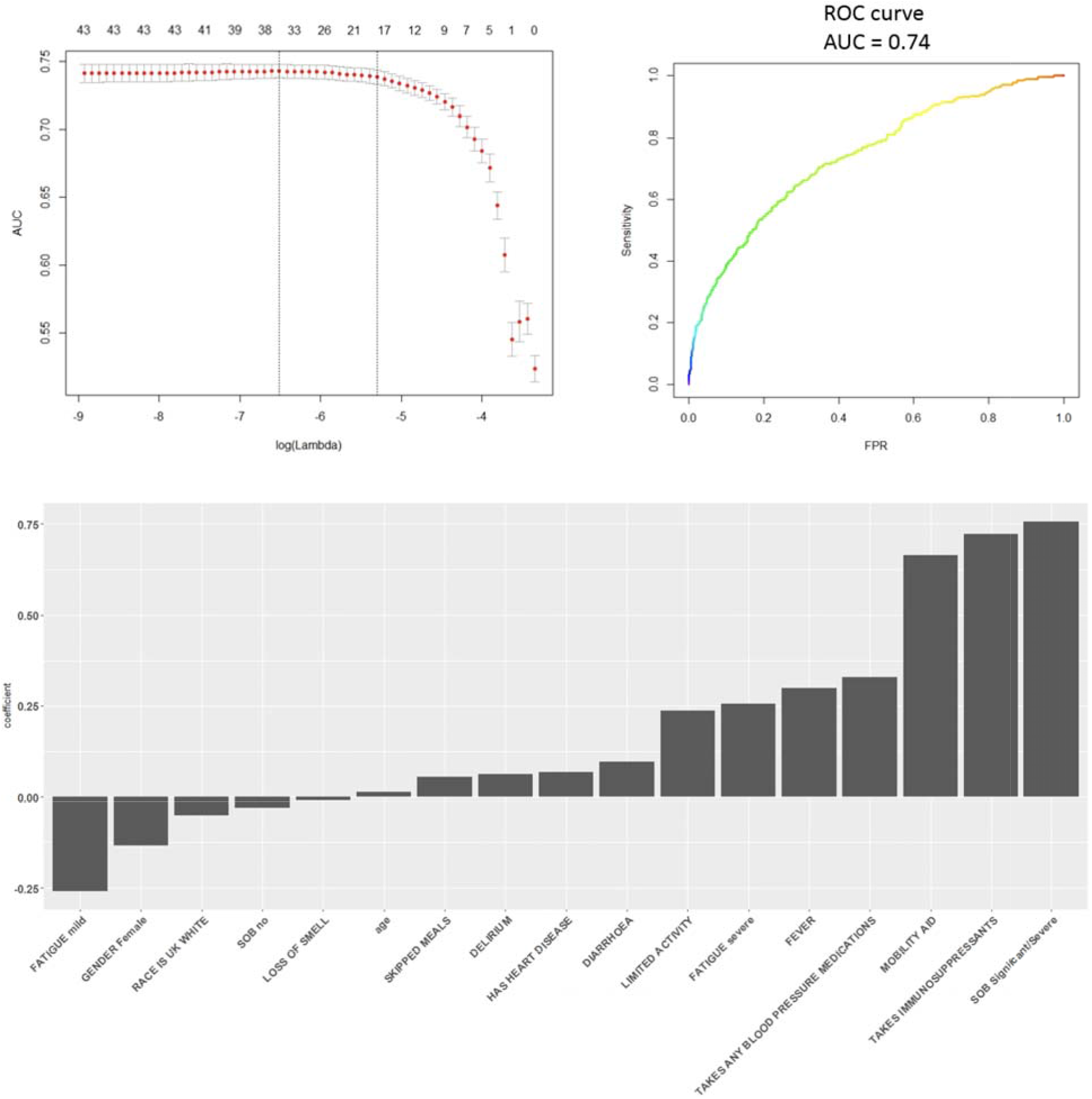
Results of Elastic Net Regression using scenario 2. Scenario 2 where for each feature, if a user indicated he/she had that feature in any of his/her entire entries, we labeled that feature as positive for that user. (A) The performance in terms of cross-validation area under the Reciever Operating Curve (AUC) for validated Elastic Net Regression on the training set across different values of lambda. (B) The AUC of the trained Elastic Net model applied on a holdout test dataset. (C) The most important features selected by the Elastic Net model. Negative coefficients indicate a negative association with outcome and vice versa. Features selected are similar to scenario 1. Predictive performances are also comparable.

**Supplementary Figure 7.**
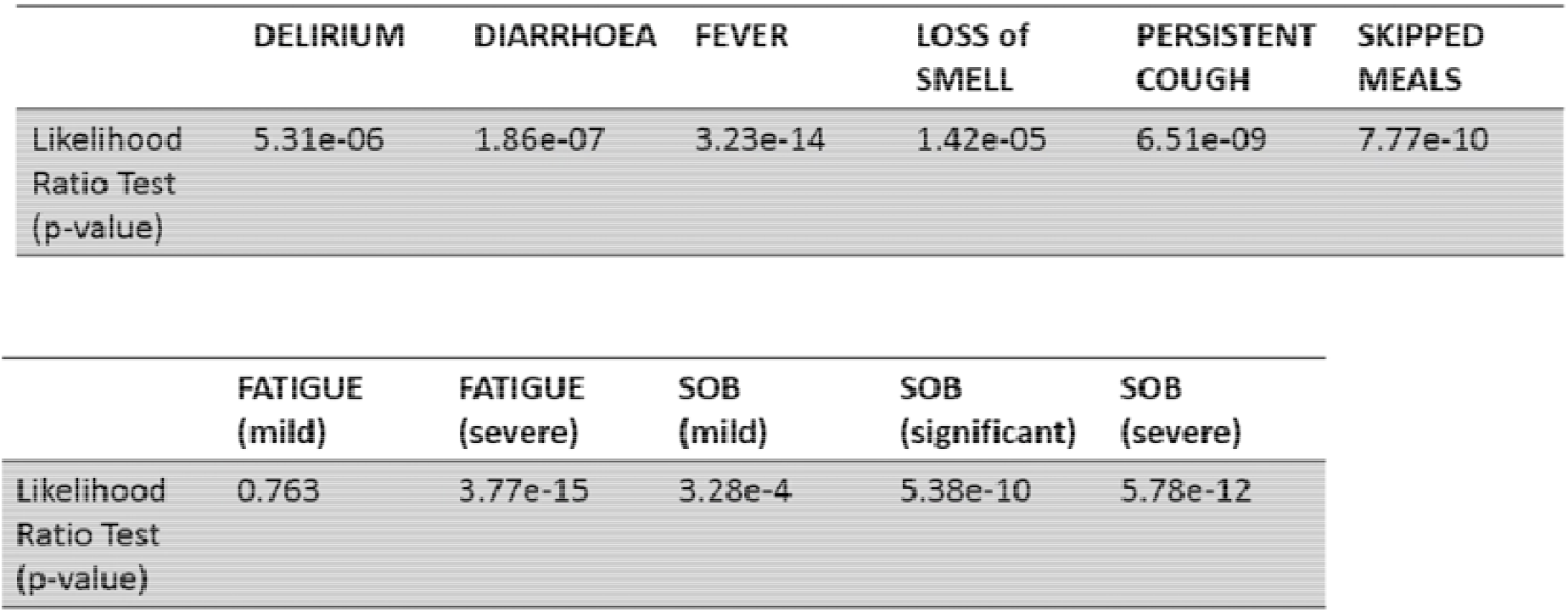
Likelihood ratio test between positive and negative groups. A 20 days window was examined for positive and negative cases. For each day, the frequency of users having the feature for the positive and negative groups is plotted. Linear regression where the frequency is regressed on the days before the last day. Slope and intercepts were obtained and the likelihood ratio test was used to evaluate whether the slopes were statistically different. P-value < 0.05 indicates the positive and negative groups have statistically different slopes.

## Acknowledgements

This work uses data provided by participants of the COVID-19 Symptoms Study, developed by ZOE Global Limited with scientific and clinical input from King’s College London. We would also like to acknowledge all data providers who made anonymised data available for research.

We wish to acknowledge the collaborative partnership that enabled acquisition and access to the de-identified data, which led to this output. The collaboration was led by BREATHE – The Health Data Research Hub for Respiratory Health, in partnership with SAIL Databank. We wish to acknowledge the input of ZOE Global Limited and King’s College London in their development and sharing of the data, and their input into the understanding and contextualisation of data for COVID-19 research. All research conducted has been completed under the permission and approval of SAIL independent Information Governance Review Panel (IGRP) project number 1088, project lead Dr Dina Radenkovic.

